# Ultrasonic Cardiac Output Monitor-Guided Fluid Resuscitation in Adult Diabetic Ketoacidosis: A Randomised Controlled Trial

**DOI:** 10.1101/2025.09.06.25322297

**Authors:** Yan Bo

## Abstract

**BACKGROUND:** Diabetic ketoacidosis (DKA) is a life-threatening complication of diabetes mellitus, requiring prompt and effective fluid resuscitation. Current guidelines rely on empirical fluid protocols that fail to incorporate real-time haemodynamic monitoring, potentially leading to fluid overload or insufficient resuscitation.

**Aims:** To evaluate whether non-invasive haemodynamic monitoring with the Ultrasonic Cardiac Output Monitor (USCOM) combined with passive leg raising (PLR) optimises fluid resuscitation in adults with diabetic ketoacidosis (DKA) without impairing metabolic recovery.

**Methods:** In this single-centre, single-blind, randomised controlled pilot trial, adults with DKA were allocated (1:1) to USCOM-guided resuscitation or sham USCOM monitoring. In the intervention group, additional fluid boluses (250 mL crystalloids over 15 min) were administered only when stroke volume increased by ≥10% during PLR. The primary outcome was net fluid balance at 24, 48, and 72 hours. Secondary outcomes included urine output, length of hospital stay, glycaemic control, urinary ketone clearance, and adverse events.

**Results:** Fifty participants (mean age 48.0 ± 16.2 years; 42% male) were enrolled. Compared with controls, the USCOM group demonstrated significantly lower net fluid balance at 48 hours (−1246.8 mL; P = 0.007) and 72 hours (−1759.5 mL; P = 0.001), driven by greater urine output in the first 24 hours (+952.6 mL; P = 0.002). Length of hospital stay was reduced by 1.92 days (P = 0.034). No significant differences were observed in blood glucose trajectories, urinary ketone clearance, or adverse event rates.

**Conclusions:** USCOM-guided resuscitation in adult DKA reduces net fluid accumulation and shortens hospitalisation without compromising metabolic recovery. This dynamic, non-invasive strategy offers a feasible approach to precision fluid management, with potential applicability in diverse healthcare settings.

**Trial registration:** ChiCTR2500103388.

## 1 Introduction

Diabetic ketoacidosis (DKA), a life-threatening complication of diabetes mellitus, is characterised by hyperglycaemia, metabolic acidosis, and ketonaemia. Prompt intervention is essential to prevent multi-organ failure and death (1,2). Current clinical guidelines lack standardisation for calculating fluid resuscitation volumes and determining optimal fluid resuscitation timing in DKA, creating uncertainty in clinical decision-making (2,3). Although fluid resuscitation remains fundamental to DKA management, clinicians face a critical dilemma: excessive fluid administration increases risks of cerebral or pulmonary oedema, whereas restrictive strategies may prolong hypoperfusion and delay metabolic recovery (4). Existing guidelines rely on static parameters to guide fluid therapy; however, these metrics poorly reflect true volume status in hyperosmolar states (2). Consequently, suboptimal fluid management contributes to iatrogenic complications in up to 30% of DKA patients (5,6)

Conventional invasive monitoring techniques deliver precise haemodynamic data but are limited by procedural risks and high costs (7). Emerging non-invasive alternatives such as passive leg raising (PLR) and pulse pressure variation (PPV) offer practical solutions; however, their accuracy diminishes in spontaneously breathing patients or those with arrhythmias (8). The Ultrasonic Cardiac Output Monitor (USCOM) addresses these limitations by integrating Doppler-based aortic flow measurements with real-time haemodynamic analytics (9). Unlike traditional echocardiography requiring specialised training, USCOM enables rapid assessments within three minutes with minimal operator dependency (10). Its portability and cost-effectiveness render it particularly suitable for emergency settings (9).

Despite promising applications in postoperative care (11), USCOM’s role in DKA fluid resuscitation remains underexplored. Existing studies are constrained by small sample sizes and lack of protocol standardisation. This randomised controlled trial tests the hypothesis that USCOM-guided fluid therapy using a ΔSV ≥10% threshold via PLR reduces fluid overload while maintaining metabolic control in DKA. By integrating dynamic haemodynamic monitoring into resuscitation protocols, we aim to establish an evidence-based strategy balancing efficacy and safety in acute metabolic crises. Our work extends the investigation by Kuppermann et al. on fluid infusion rates and adverse outcomes in paediatric DKA (4).

The primary objective of this study is to determine whether USCOM-guided fluid management improves clinical outcomes in DKA. Through a randomised controlled design, we evaluate the safety and efficacy of USCOM-assisted resuscitation, addressing the longstanding clinical question: how can clinicians identify the minimal effective fluid volume or optimal cessation point during DKA resuscitation?

## 2 Methods

### 2.1 Patient and public involvement

Patients and the public were not involved in the design, conduct and reporting of the trial.

### 2.2 Trial setting

This was a two-arm, 1-to-1 randomised parallel-controlled, single-blind, pilot clinical trial.

This pilot trial was conducted in Jiangsu Province, China. Participants were enrolled from the Emergency Department of Yangzhou University Affiliated Hospital. The study complied with the Declaration of Helsinki and received approval from the hospital’s Institutional Review Board (IRB-06-004). All participants provided written informed consent prior to enrollment. The protocol was registered with the Chinese Clinical Trial Registry (ChiCTR2500103388) and conducted in accordance with CONSORT guidelines (12).

### 2.3 Eligibility criteria

Participants meeting all inclusion criteria were enrolled in this pilot trial. Those subsequently found to meet exclusion criteria during comprehensive evaluation were advised to withdraw voluntarily with explanation. DKA diagnosis required fulfillment of both ICD-10 criteria and the Chinese Guidelines for Diagnosis and Treatment of Hyperglycemic Crises (13).

Inclusion criteria: (1) aged ≥18 years; (2) current diagnosis of diabetic ketoacidosis; (3) laboratory confirmation: blood glucose >13.9 mmol/L, serum ketones ≥3 mmol/L or positive urine ketones, arterial pH <7.3 and/or bicarbonate <18 mmol/L; (4) systolic blood pressure ≥90 mmHg without vasopressor support

Exclusion criteria: Presence of conditions contraindicating non-invasive ultrasound assessment. (1) cardiopulmonary comorbidities (class III-IV heart failure, severe COPD);(2) structural valvular abnormalities (aortic/pulmonary stenosis or regurgitation); (3) thoracic deformities;(4) history of thoracic surgery

All fluid resuscitation procedures were administered by attending emergency physicians within designated treatment bays of the Emergency Department.

### 2.4 Intervention and comparator

All participants received standardized fluid resuscitation per the 2022 Chinese Consensus Guidelines for Diabetic Ketoacidosis and Hyperosmolar Hyperglycemic State (13), with adjustments based on vital signs and laboratory parameters.

The control group device was engineered to physically resemble the USCOM 1A (USCOM Ltd, Australia) but displayed randomly generated hemodynamic values. Both devices maintained identical user interfaces and operational workflows to ensure participant blinding.

Intervention group participants underwent hemodynamic assessment using USCOM 1A with a 2.2 MHz transducer, following Phillips et al.’s standardized protocol (14).

The primary procedure consisted of passive leg raising (PLR) testing and haemodynamic measurements. During the PLR test, patients transitioned from a 45° semi-recumbent position to a supine position with the leg elevated at 45° for 2 minutes.A meta-analysis by Monnet X et al. showed that the overall correlation between plr-induced changes in cardiac output (CO) and fluid-induced changes in CO was 0.76 (0.73 - 0.80). For plr-induced CO changes, the combined sensitivity was 0.85 (0.81 ∼ 0.88), the combined specificity was 0.91 (0.88 ∼ 0.93), and the area under the ROC curve was 0.95 ± 0.01. The optimal threshold was a plr-induced CO elevation of ≥ 10 ± 2% (15). A sensitivity of 85% for PLR supports its applicability in spontaneously breathing patients. Haemodynamic measurements were performed by measuring the aortic velocity-time integral (VTI) in the left parasternal window. The formulae for calculating cerebral stroke volume (SV) and cardiac output (CO) were SV = VTI × aortic valve area (cm²) and CO = SV × heart rate.

The fluid response threshold was determined as follows: ΔSV ≥10% after PLR triggering an additional fluid dose (250 mL of crystal over 15 minutes); ΔSV <10% suggests flow restriction. We adopted ΔSV ≥10% as the fluid response threshold proposed by Marik PE et al. to improve the reliability of the study (16).

Participants assigned to receive USCOM monitoring feedback were required to determine whether the fluid resuscitation threshold was reached by readings from the PLS and USCOM devices prior to each fluid resuscitation. If the participant did not reach the fluid resuscitation threshold, fluid resuscitation was started, otherwise fluid resuscitation was paused to continue observation. Participants who were assigned not to receive USCOM monitoring feedback did not make a fluid resuscitation threshold judgement before each fluid resuscitation, and were rehydrated directly until the fluid resuscitation was complete.

Diabetic ketoacidosis and hyperglycaemic hyperosmolar state received standard fluid resuscitation according to the Chinese clinical practice consensus guideline 2022 and was adjusted according to vital signs and laboratory parameters (13).

### 2.5 Outcomes

The primary outcome was prespecified as the amount of fluid resuscitation (current fluid resuscitation, net fluid resuscitation, and urinary output). Net fluid intake represents the difference between the participant’s current total fluid resuscitation and urinary output. In the study outcomes, we set 24 hours, 48 hours, and 72 hours as time points to describe fluid resuscitation volume.

Secondary outcome indicators were pre-specified as the incidence of complications, urinary ketone clearance, blood glucose values, technical parameters of the USCOM, and days of hospitalisation. Complications denote clinical adverse events, whether or not related to treatment, that occurred after the participant received either the intervention group or the comparison group up until the time of discharge. We only counted the types of complications that occurred in the participants, regardless of the frequency.The technical parameters of the USCOM were pre-specified in terms of stroke volume (SV) and cardiac output (CO).The technical parameters of the USCOM were only required to be fitted to the participant, and the screen of the device would display the specific values. In the results of the study, only participants in the intervention group had the technical parameters of the USCOM measured; for the baseline data, we used the USCOM to measure the participants before they received their first fluid resuscitation solution. We measured the technical parameters of the USCOM each time a participant received fluid resuscitation. Urinary ketone clearance and blood glucose values were confirmed by laboratory tests based on participants’ venous blood samples. The time points for reporting of blood glucose values in the study results were the first laboratory examination of the venous blood sample after check-in, and 24 hours after admission and 72 hours after admission. Urinary ketone clearance was only reported in the study results corrected for the day of the participant’s DKA (the time point when the last fluid resuscitation therapy was completed).

These outcome indicators are expressed using mean and standard deviation if they follow a normal distribution, and median and quartiles otherwise. The time points for measurement and reporting of outcomes are shown in the **Supplementary document**.

### 2.6 Harms

The non-invasive ultrasound device is a non-invasive monitoring instrument and therefore does not pose a hazard to participants.

### 2.7 Sample size

A 30% reduction in fluid intake at 48 hours was assumed [(effect values d = 0.8, α = 0.05, β = 0.20, σ = 1,834 mL, Δ = 1,247 mL (expected difference)]. At 20% attrition, 25 patients were required in each group.

The sample size was calculated using the formula:

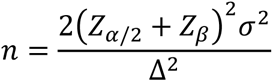

### 2.8 Randomisation and blinding

Yi-fei Chen, a practising emergency physician, used a computer to generate random numbers between 0 and 1, and subsequently telephoned doctors involved in the trial to inform them of the allocation results. Participants who received a random number between 0 and 0.5 (the top 50%) would be assigned to the intervention group, while those between 0.5 and 1 (the bottom 50%) would be assigned to the control group. As recruitment progresses, either group will reach the recruitment endpoint first, i.e. 25 participants who are eligible for the study and do not meet the exclusion criteria are recruited. This safeguards that participants are randomised 1:1 to the intervention and control groups.

The grouping and intervention method that the participant receives is unknown only to the participant. This single-blind approach we realised in two ways. First, a similar non-invasive ultrasound monitoring device was used as a control, which was not actually functional. Second, participants were not informed of the expected desired outcome. Whether the amount of fluid resuscitation therapy received by the participant during the period just reached the fluid resuscitation threshold was known only to the emergency physician. The emergency physician responsible for fluid management was aware of the groupings, but the data analyst was blinded to the groupings.

Second, in order to ensure that the participants in the two groups were using similar devices, participants in the control group would use a sham USCOM device.

### 2.9 Statistical methods

USCOM measurements were performed by two trained emergency physicians, averaging three readings to minimise operator variation. Laboratory parameters (pH, bicarbonate, ketones) were analysed by a blinded technician using standardised assays.

All clinical data were analysed by a data difference test to assess differences in the effect of USCOM monitoring feedback between the intervention and control groups. Statistical tests were two-sided and performed at the 0.05 level.

Data were analysed using SPSS 27.0 (IBM Corp.). Normally distributed continuous variables were expressed as mean±SD and compared by independent t tests. Non-normal data were expressed as median (IQR) and analysed by Mann-Whitney U test. Categorical variables were expressed as frequencies (%) and compared using χ^2^ or Fisher exact tests.

Predefined subgroup analyses were stratified by age of participants, with 60 years as the cut-off point.

## 3 Results

### 3.1 Participant flow

From 6 March 2022 until 1 August 2025, we initiated this pilot clinical trial and a total of 69 participants were assessed for study eligibility. During the assessment of study eligibility, 12 participants declined to sign the informed consent form, 4 participants declined to participate due to heart valve pathology, and 3 participants declined to participate due to haemodynamic instability. The remaining 50 participants were randomised 1 to 1 to the USCOM monitoring group (intervention group) and the sham USCOM monitoring group (control group). No participants withdrew during the 14-day treatment and follow-up period. All 50 participants completed the planned study protocol. Data from all participants were included in the analysis (**Supplementary document**). We summarized the research design and findings using graphical abstract (**Figure 1**).

**Figure 1.**
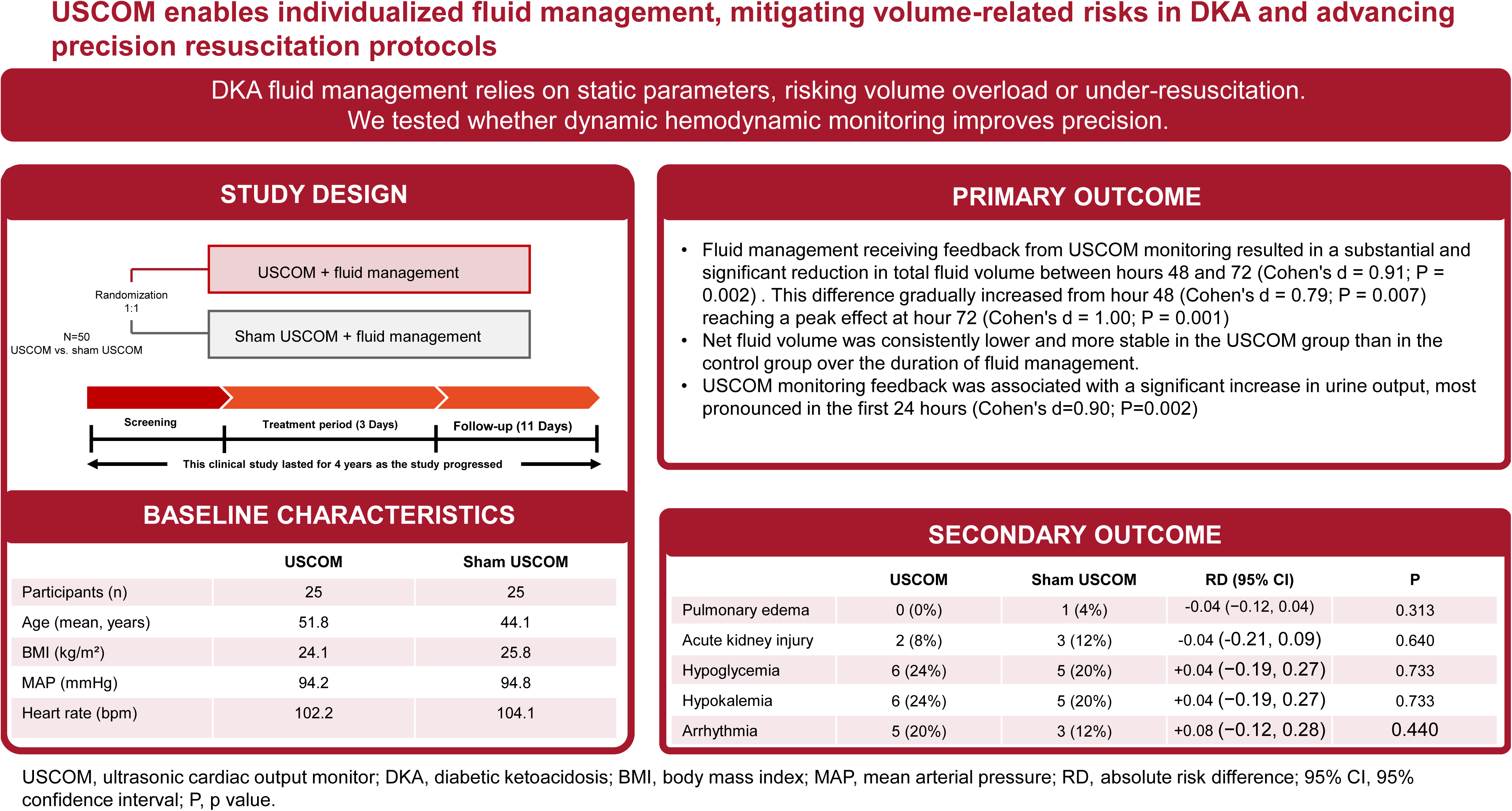
Graphical abstract. This section drawn by Figdraw provides a concise overview of the participants’ study design, enrollment procedures, and baseline characteristics, while also presenting the primary and secondary outcomes of this clinical trial.

### 3.2 Baseline data

Baseline demographics and clinical parameters were balanced between the groups (**Supplementary document**). The difference in clinical baseline data between the two groups was not statistically significant. This implies that the treatment data of the participants in these two groups are of comparative value.

### 3.3 Efficacy Analyses

#### 3.3.1 Primary Endpoints

Participants in both groups achieved DKA resolution within 3 days of fluid resuscitation. This was consistent with the current clinical practice. Reduced fluid volumes correlate with faster recovery without compromising treatment efficacy, we analyzed cumulative fluid resuscitation volumes between groups (**Table 1**).

**Table 1.** Fluid in USCOM-Guided vs. Control Groups.

| Time Point | USCOM Group<br>(n=25) | Control Group<br>(n=25) | t-value | Cohen's d | Mean Difference | 95%CI | P-value |
| --- | --- | --- | --- | --- | --- | --- | --- |
| **Fluid replenishment (mL)** |  |  |  |  |  |  |  |
| 24h | 6514.04 ± 1886.14 | 6230.8 ± 1404.68 | 0.60 | 0.17 | 283.24 | (-662.66, 1229.14) | 0.550 |
| 48h | 3052.68 ± 914.64 | 3010.96 ± 1358.32 | 0.13 | 0.04 | 41.72 | (-619.18, 702.62) | 0.899 |
| 72h | 1751.04 ± 533.62 | 2328.88 ± 719.03 | 3.23 | 0.91 | -577.84 | (217.74, 937.94) | **0.002** |
| Total 72h | 11317.76 ± 2436.41 | 11570.64 ± 2726.75 | 0.35 | 0.10 | -252.88 | (-1217.82, 1723.58) | 0.730 |
| **Net fluid replenishment (mL)** |  |  |  |  |  |  |  |
| 24h | 2681.68 ± 1864.92 | 3351.00 ± 1208.02 | 1.51 | 0.44 | -669.32 | (-228.37, 1567.01) | 0.136 |
| 48h | 2934.12 ± 1498.71 | 4180.96 ± 1666.60 | 2.78 | 0.79 | -1246.84 | (345.37, 2148.31) | **0.007** |
| 72h | 2916.36 ± 1511.30 | 4675.84 ± 1981.00 | 3.53 | 1.00 | -1759.48 | (757.42, 2761.54) | **0.001** |
| **Urine output (mL)** |  |  |  |  |  |  |  |
| 24h | 3832.36 ± 994.37 | 2879.80 ± 1125.67 | 3.17 | 0.90 | 952.56 | (348.48, 1556.64) | **0.002** |
| 48h | 2800.24 ± 1383.00 | 2181.00 ± 1089.24 | 1.76 | 0.50 | 619.24 | (-89.03, 1327.51) | 0.084 |
| 72h | 1768.8 ± 822.27 | 1834.00 ± 958.83 | 0.26 | 0.07 | -65.20 | (-442.84, 573.24) | 0.795 |

24 hours indicates continuous time counted from the time of the first rehydration to within the 24th hour. 48 hours indicates continuous time counted from the time of the first rehydration to within the 24th hour to within the 48th hour. 72 hours indicates continuous time counted from the time of the first rehydration to within the 48th hour to within the 72nd hour. Cumulative 72 hours indicates continuous time from the time of the first rehydration to within the 72nd hour. 95% CI indicates 95% confidence interval. P values less than 0.05 are bolded to indicate significance.

Contrary to our initial hypothesis, the intervention group receiving USCOM-guided resuscitation received higher total fluid volumes than controls during the initial phase. This suggests that USCOM-guided management did not restrict overall fluid administration. However, a significant reversal emerged between 48–72 hours, where the intervention group received substantially less fluid (Cohen’s d=0.91; P=0.002). This divergence increased progressively from 48 hours (Cohen’s d=0.79; P=0.007) to a peak effect at 72 hours (Cohen’s d=1.00; P=0.001). Crucially, net fluid balance (total input minus urinary output) was consistently lower in the USCOM group throughout resuscitation (**Table 1**).

To elucidate the mechanism underlying the reduced net fluid balance with USCOM guidance [mean reduction: 1759.48 mL (95% CI: 757.42–2761.54 mL); P=0.001], we analyzed urinary output. USCOM guidance was associated with a significantly higher urine volume, most pronounced in the first 24 hours (Cohen’s d=0.90; P=0.002), followed by gradual attenuation. This demonstrates that USCOM facilitates enhanced diuresis during active resuscitation, particularly within the first 48 hours. Consequently, despite comparable total fluid input early on, USCOM guidance reduced net fluid accumulation by promoting greater urine output.

Excessive fluid administration beyond individual tolerance during DKA resuscitation increases the risk of adverse events (volume overload), prolonging hospitalization. USCOM-guided management mitigates this risk by dynamically optimizing fluid delivery, thereby reducing net fluid retention while ensuring effective acidosis correction.

#### 3.3.2 Secondary Endpoints

Both groups received identical diabetes management protocols. Dr. Bo postulated that USCOM-guided fluid resuscitation might potentially disrupt glucose metabolism via electrolyte alterations, thereby compromising glycemic control. However, comparative analysis of venous blood glucose levels during resuscitation revealed no significant intergroup differences (**Table 2**). This indicates that reducing net fluid balance through enhanced diuresis under USCOM guidance does not impede metabolic recovery or hyperglycemia management.

**Table 2.** Laboratory Examination in USCOM-Guided vs. Control Groups.

| Time Point | USCOM Group<br>(n=25) | Control Group<br>(n=25) | t-value | Cohen's d | Mean Difference | 95%CI | P-value |
| --- | --- | --- | --- | --- | --- | --- | --- |
| Blood Glucose (mmol/L) |  |  |  |  |  |  |  |
| 24h | 16.07±3.62 | 15.99±4.51 | 0.07 | 0.02 | 0.08 | (-2.25, 2.41) | 0.944 |
| 48h | 15.81±3.54 | 14.84±4.15 | 0.89 | 0.25 | 0.97 | (-1.23, 3.17) | 0.377 |
| 72h | 13.85±4.09 | 13.32±3.23 | 0.51 | 0.14 | 0.53 | (-1.56, 2.62) | 0.612 |
| Time for urinary ketone bodies to turn negative (day) |  |  |  |  |  |  |  |
| Discharge | 6.04±2.29 | 6.28±2.59 | 0.35 | 0.10 | -0.24 | (-1.15, 1.63) | 0.730 |
| Length of hospitalization (day) |  |  |  |  |  |  |  |
| Discharge | 7.8±2.77 | 9.72±3.38 | 2.18 | 0.62 | -1.92 | (0.15, 3.69) | 0.034 |
| Hemodynamic Parameters |  |  |  |  |  |  |  |
| SV (mL) |  |  |  |  |  |  |  |
| Baseline | 29.8±4.5 |  | 14.82 | 2.96 | 38.8 | (33.2, 44.4) | <0.001 |
| Post-Resuscitation | 68.6±9.7 |  |  |  |  |  |  |
| CO (L/min) |  |  |  |  |  |  |  |
| Baseline | 3.0±0.7 |  | 12.00 | 2.40 | 2.4 | (1.9, 2.9) | <0.001 |
| Post-Resuscitation | 5.4±1.0 |  |  |  |  |  |  |

24 hours indicates continuous time counted from the time of the first rehydration to within the 24th hour. 48 hours indicates continuous time counted from the time of the first rehydration to within the 24th hour to within the 48th hour. 72 hours indicates continuous time counted from the time of the first rehydration to within the 48th hour to within the 72nd hour. Cumulative 72 hours indicates continuous time from the time of first rehydration to within the 72nd hour. 95% CI indicates 95% confidence interval. P values less than 0.05 are bolded to indicate significance. Discharge indicates the point at which the participant was discharged. Urinary ketone turnaround time is the cumulative number of hospital days assessed daily until the last assessment was a negative result. SV indicates Stroke Volume. CO indicates Cardiac Output. t-tests of SV and CO are paired t-tests because they are more sensitive to eliminating individual patient differences when comparing the same patient before and after treatment. Baseline indicates data measured before the first rehydration therapy. Post-rehydration therapy indicates data measured before the last rehydration therapy.

This finding receives secondary validation from the comparable time to urinary ketone clearance between groups (P = 0.730).

Regarding hospitalization duration, US data report a mean hospital stay of 3.5 days for DKA resolution. Following consultation between Dr. Bo and Prof. Dr. Chen, we interpret this US benchmark as reflecting acidosis correction alone, excluding management of complications or underlying comorbidities. By contrast, clinical practice patterns in China typically involve extended hospitalization until resolution of concomitant issues.

USCOM-guided management reduced mean hospitalization by 1.92 days compared to controls (Cohen’s d = 0.62; P = 0.034) (**Table 2**). This substantial reduction suggests that USCOM implementation can shorten hospital stays for DKA patients in the Chinese healthcare context, thereby alleviating system burdens.

### 3.4 Safety

Despite the non-invasive nature of USCOM, we systematically assessed complications in both groups during DKA resuscitation, irrespective of potential treatment association, to capture all safety events. As shown in **Supplementary document and Figure 1**, USCOM-guided resuscitation showed a non-significant numerical increase in the absolute risk difference for overall complications compared to controls (Cohen’s d=0.274; P=0.440). Although complication rates did not differ statistically, we further analyzed hemodynamic parameters to explore this unexpected finding.

Hemodynamic status significantly improved in the USCOM-guided group between initial and final resuscitation assessments (**Table 2**). Both stroke volume (SV) and cardiac output (CO) demonstrated substantial increases, with effect sizes exceeding 2.0, indicating profound hemodynamic enhancement following USCOM-guided management. We hypothesize that this rapid, significant hemodynamic improvement over the 3-day resuscitation period may be associated with the observed non-significant trend toward increased complications.

### 3.5 Ancillary analyses

Pre-specified subgroup analysis stratified by age (cutoff: 60 years) confirmed the overall trend of reduced fluid load with USCOM guidance.

In patients <60 years, USCOM-guided resuscitation stabilized net fluid balance at approximately 2796 mL per 24 hours (**Table 3**).

**Table 3.** Subgroup analysis with 60 years old as the dividing point: Fluid in USCOM-Guided vs. Control Groups.

| Time Point | USCOM Group | Control Group | t-value | Cohen's d | Mean Difference | 95%CI | P-value |
| --- | --- | --- | --- | --- | --- | --- | --- |
| **Less than 60 Ages [USCOM Group (N=17) vs Control Group (N=21)]** |  |  |  |  |  |  |  |
| ● Fluid replenishment (mL) |  |  |  |  |  |  |  |
| 24h | 6637.35±2220.25 | 6232.43±1402.32 | 0.69 | 0.22 | 404.92 | (-794.0, 1603.8) | 0.497 |
| 48h | 3035.59±921.23 | 3141.10±1403.68 | -0.27 | 0.09 | -105.51 | (-907.6, 696.6) | 0.787 |
| 72h | 1727.76±456.78 | 2300±699.01 | -2.91 | 0.95 | -572.24 | (-973.1, -171.3) | **0.006** |
| Total 72h | 11400.71±2677.67 | 11669.24±2840.30 | -0.30 | 0.10 | -268.53 | (-2079.5, 1542.5) | 0.773 |
| ● Net fluid replenishment (mL) |  |  |  |  |  |  |  |
| 24h | 2759.12±2007.14 | 3200.76±1081.40 | -0.82 | 0.27 | -441.64 | (-1555.0, 671.7) | 0.424 |
| 48h | 2748.94±1400.17 | 4036.62±1816.50 | -2.40 | 0.78 | -1287.68 | (-2376.0, -199.4) | **0.021** |
| 72h | 2796.71±1579.76 | 4442.81±2007.73 | -2.76 | 0.89 | -1646.10 | (-2855.7, -436.5) | **0.009** |
| ● Urine output (mL) |  |  |  |  |  |  |  |
| 24h | 3878.24±1083.97 | 3031.67±1144.61 | 2.32 | 0.76 | 846.57 | (108.7, 1584.4) | **0.026** |
| 48h | 3045.76±1584.61 | 2305.24±1134.93 | 1.68 | 0.54 | 740.52 | (-153.2, 1634.2) | 0.102 |
| 72h | 1680±635.42 | 1899.05±1016.13 | -0.81 | 0.25 | -219.05 | (-768.5, 330.4) | 0.423 |
| **More than 60 Ages [USCOM Group (N=8) vs Control Group (N=4)]** |  |  |  |  |  |  |  |
| ● Fluid replenishment (mL) |  |  |  |  |  |  |  |
| 24h | 6252±734.71 | 6222.25±1417.23 | 0.05 | 0.03 | 29.75 | (-1274.9, 1334.4) | 0.961 |
| 48h | 3089±899.41 | 2327.75±791.66 | 1.43 | 0.87 | 761.25 | (-372.2, 1894.7) | 0.184 |
| 72h | 1800.5±665.55 | 2503±793.00 | -1.63 | 1.00 | -702.5 | (-1623.8, 218.8) | 0.135 |
| Total 72h | 11141.5±1807.49 | 11053±1952.32 | 0.08 | 0.05 | 88.5 | (-2342.9, 2520.0) | 0.939 |
| ● Net fluid replenishment (mL) |  |  |  |  |  |  |  |
| 24h | 2517.125±1217.20 | 4139.75±1496.80 | -2.04 | 1.24 | -1622.625 | (-3286.0, 39.7) | 0.069 |
| 48h | 3327.625±1620.86 | 4938.75±1473.68 | -1.67 | 1.02 | -1611.125 | (-3686.8, 464.5) | 0.127 |
| 72h | 3170.625±1318.32 | 5899.25±1246.86 | -3.44 | 2.10 | -2728.625 | (-4385.3, -1071.9) | **0.005** |
| ● Urine output (mL) |  |  |  |  |  |  |  |
| 24h | 3734.875±761.75 | 2082.5±517.27 | 3.87 | 2.37 | 1652.375 | (701.1, 2603.7) | **0.002** |
| 48h | 2278.5±491.07 | 1528.75±395.61 | 2.64 | 1.61 | 749.75 | (116.4, 1383.2) | **0.024** |
| 72h | 1957.5±1096.61 | 1542.5±474.26 | 0.71 | 0.43 | 415 | (-884.3, 1714.3) | 0.495 |

24 hours indicates continuous time counted from the time of the first rehydration to within the 24th hour. 48 hours indicates continuous time counted from the time of the first rehydration to within the 24th hour to within the 48th hour. 72 hours indicates continuous time counted from the time of the first rehydration to within the 48th hour to within the 72nd hour. Cumulative 72 hours indicates continuous time from the time of the first rehydration to within the 72nd hour. 95% CI indicates 95% confidence interval. P values less than 0.05 are bolded to indicate significance.

A similar benefit was observed in patients ≥60 years (**Table 3**), suggesting particular relevance for those with comorbid cardiac or renal insufficiency, where minimizing net positive fluid balance reduces complication risks associated with volume overload.

## 4 Discussion

### 4.1 Research significance

Our findings address critical gaps in DKA fluid management identified by landmark studies.

Umpierrez and Korytkowski (2016) clearly state that fluid resuscitation is the first step in the treatment of DKA, aimed at correcting dehydration and restoring circulatory volume and renal function. The average dehydration is approximately 100 ml/kg in adult DKA patients and 70 ml/kg in children, but clinical assessment of the degree of dehydration often relies on subjective signs with limited accuracy (17). Over-hydration may lead to cerebral and pulmonary oedema, while under-hydration delays correction of metabolic disturbances. At this time, fluid management mainly relies on the empirical formula proposed by Holliday-Segar, and the lack of real-time haemodynamic monitoring makes it difficult to accurately adjust the rate and volume of rehydration.

A 2018 study based on the DKA rehydration debate in children noted no significant difference in the risk of cerebral oedema between rapid (50% rehydration within 4 hours) and slow (48-hour rehydration) rehydration regimens for DKA in children, but emphasised the need for dynamic adjustments based on blood sodium and osmolality (3). However, the guidelines still recommend the empirical strategy of ‘deficit + maintenance by body weight’, which lacks a direct assessment of cardiac function and blood volume status, especially in patients with cardiac insufficiency or shock, and may lead to complications due to volume overload.

A systematic review by Dhatariya et al. (2020) showed that the prevalence of electrolyte disturbances and haemodynamic instability in patients with DKA is as high as 30-60%, and that conventional monitoring is less sensitive to early volume overload or underload (18). This implies that fluid mismanagement is associated with prolonged hospital stay and multi-organ dysfunction, highlighting the need for real-time haemodynamic monitoring, but existing guidelines do not recommend specific bedside monitoring tools.

Umapathysivam et al., (2024) reported that normoglycaemic DKA induced by SGLT2 inhibitors in non-diabetic patients is often accompanied by cardiac insufficiency, and that rehydration needs to be balanced between volume resuscitation and the risk of heart failure, which is difficult to apply with traditional weight-based protocols (19). This report implies that patients with DKA are more sensitive to volume changes and require precise assessment of cardiac output and intravascular volume, and that existing tools are not suitable for rapid bedside adjustments.

These points from highly cited literature illustrate that USCOM, as a non-invasive ultrasound cardiac output monitoring device, measures CO and SV in real time, which directly reflects the status of blood volume and cardiac function. This provides an objective basis for DKA fluid management. In addition, by dynamically monitoring the haemodynamic parameters, the temporary physician can avoid volume overload or insufficiency caused by empirical fluid replacement, which is especially suitable for children, heart failure combined with DKA and other high-risk groups. From empirical rehydration to individualised adjustment, the core requirement of fluid management in DKA is ‘accurate assessment of volume status and cardiac function’. In this study, USCOM monitoring was introduced, and for the first time, its effectiveness in optimising rehydration strategies was verified in a randomised controlled trial, providing key evidence for the refinement and intelligence of DKA treatment.

### 4.2 Literature review

The literature review for this study was conducted under the auspices of Yan Bo, and the literature review strategy used in Yan Bo’s previous studies was adopted (20–22). The literature related to DKA rehydration studies was then reviewed according to the PRISMA guidelines (23). As of 25 March 2025, the terms ‘diabetic ketoacidosis’ and ‘rehydration’ were searched using the PubMed database. The search terms ‘diabetic ketoacidosis’ and ‘rehydration solution’ were chosen because of their relevance to the diseases investigated in this study and the purpose of the study. Existing literature using these terms was searched to identify studies that directly investigated the relationship between DKA and rehydration. We conducted a systematic search based on the methodology of Zhao et al., Ma et al. and Liang et al. (24–26). At the literature search stage, the types of study designs we were interested in included randomised controlled studies, cohort studies and retrospective observational studies. Details of this mixed study design can be found in our previous studies (21,27,28).

After a systematic search of the PubMed database, we identified 25 studies on DKA rehydration, of which 1 case report, 2 systematic evaluations, and 2 studies in which the population was clinicians were excluded. The remaining 20 studies were included in the literature review (29–48). Nine of these studies were in child populations (6040 cases), 10 were in adult populations (2322 cases), and 1 was in a colleague population that included both children and adults (20 cases), for a cumulative total of 8382 cases. In addition, we found that the proportion of children in these cases was approximately 72%, making it clear that current research on rehydration therapy for DKA focuses on children. In order to fully enhance the necessity of our study, we divided it according to the classification of the rehydration research tasks. Among these were the relationship between rehydration time and complications (n=8), choice of rehydration modality (n=9), and determination of rehydration volume and rate (n=3). The main purpose of the literature review was to use the extant literature as an external validation to test the results of our pilot clinical study. We placed all the main lessons obtained from the literature in a table (**Table 4**).

**Table 4.**
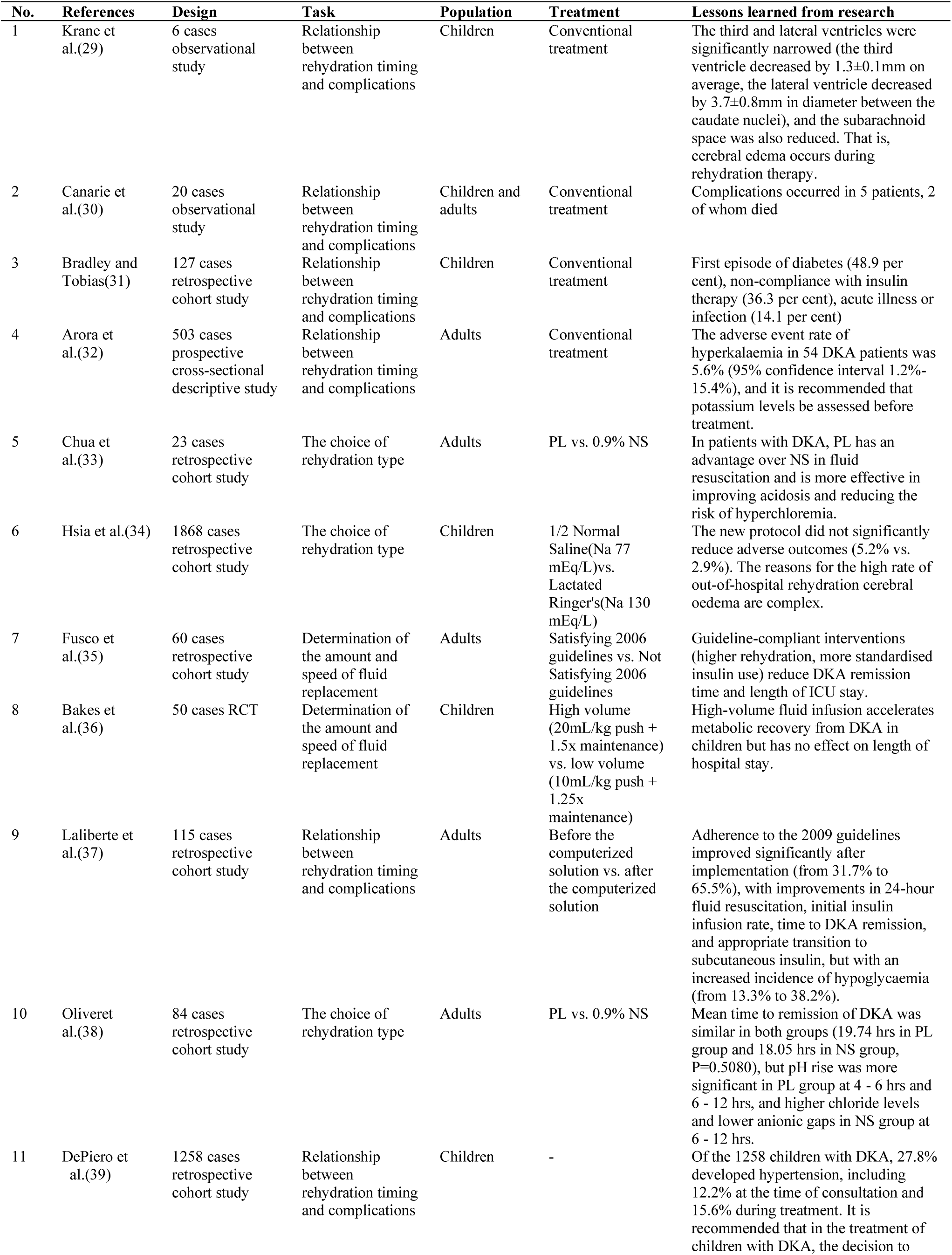
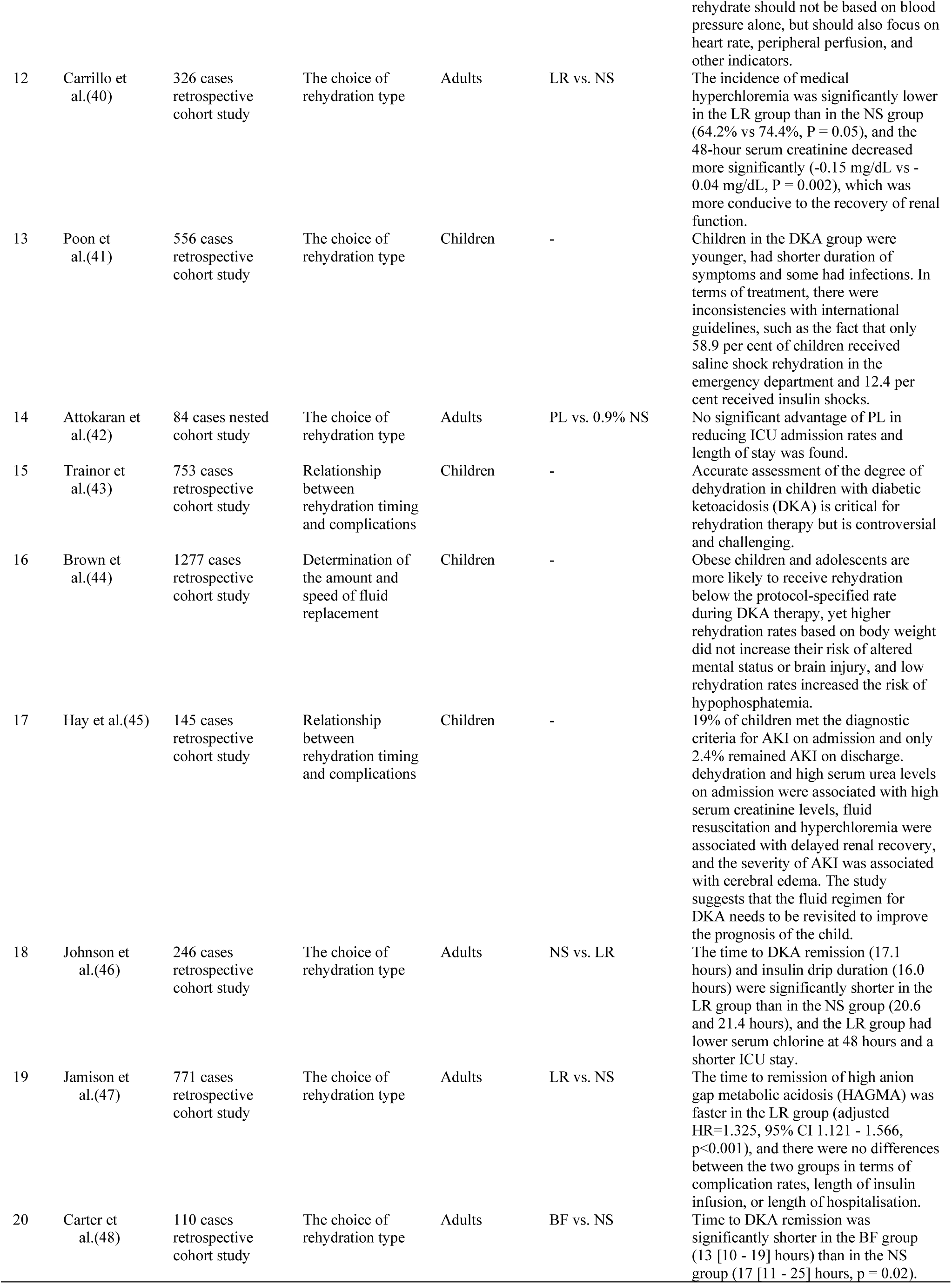
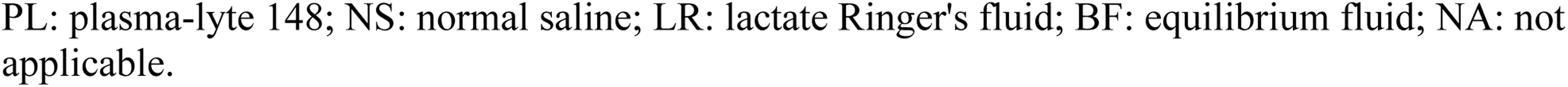
Review of 8382 cases of clinical study of DKA fluid infusion.

| No. | References | Design | Task | Population | Treatment | Lessons learned from research |
| --- | --- | --- | --- | --- | --- | --- |
| 1 | Krane et al.(29) | 6 cases observational study | Relationship between rehydration timing and complications | Children | Conventional treatment | The third and lateral ventricles were significantly narrowed (the third ventricle decreased by $1.3 \pm 0.1$ mm on average, the lateral ventricle decreased by $3.7 \pm 0.8$ mm in diameter between the caudate nuclei), and the subarachnoid space was also reduced. That is, cerebral edema occurs during rehydration therapy. |
| 2 | Canarie et al.(30) | 20 cases observational study | Relationship between rehydration timing and complications | Children and adults | Conventional treatment | Complications occurred in 5 patients, 2 of whom died |
| 3 | Bradley and Tobias(31) | 127 cases retrospective cohort study | Relationship between rehydration timing and complications | Children | Conventional treatment | First episode of diabetes (48.9 per cent), non-compliance with insulin therapy (36.3 per cent), acute illness or infection (14.1 per cent) |
| 4 | Arora et al.(32) | 503 cases prospective cross-sectional descriptive study | Relationship between rehydration timing and complications | Adults | Conventional treatment | The adverse event rate of hyperkalaemia in 54 DKA patients was 5.6% (95% confidence interval 1.2%-15.4%), and it is recommended that potassium levels be assessed before treatment. |
| 5 | Chua et al.(33) | 23 cases retrospective cohort study | The choice of rehydration type | Adults | PL vs. 0.9% NS | In patients with DKA, PL has an advantage over NS in fluid resuscitation and is more effective in improving acidosis and reducing the risk of hyperchloremia. |
| 6 | Hsia et al.(34) | 1868 cases retrospective cohort study | The choice of rehydration type | Children | 1/2 Normal Saline(Na 77 mEq/L)vs. Lactated Ringer's(Na 130 mEq/L) | The new protocol did not significantly reduce adverse outcomes (5.2% vs. 2.9%). The reasons for the high rate of out-of-hospital rehydration cerebral oedema are complex. |
| 7 | Fusco et al.(35) | 60 cases retrospective cohort study | Determination of the amount and speed of fluid replacement | Adults | Satisfying 2006 guidelines vs. Not Satisfying 2006 guidelines | Guideline-compliant interventions (higher rehydration, more standardised insulin use) reduce DKA remission time and length of ICU stay. |
| 8 | Bakes et al.(36) | 50 cases RCT | Determination of the amount and speed of fluid replacement | Children | High volume (20mL/kg push + 1.5x maintenance) vs. low volume (10mL/kg push + 1.25x maintenance) | High-volume fluid infusion accelerates metabolic recovery from DKA in children but has no effect on length of hospital stay. |
| 9 | Laliberte et al.(37) | 115 cases retrospective cohort study | Relationship between rehydration timing and complications | Adults | Before the computerized solution vs. after the computerized solution | Adherence to the 2009 guidelines improved significantly after implementation (from 31.7% to 65.5%), with improvements in 24-hour fluid resuscitation, initial insulin infusion rate, time to DKA remission, and appropriate transition to subcutaneous insulin, but with an increased incidence of hypoglycaemia (from 13.3% to 38.2%). |
| 10 | Oliveret al.(38) | 84 cases retrospective cohort study | The choice of rehydration type | Adults | PL vs. 0.9% NS | Mean time to remission of DKA was similar in both groups (19.74 hrs in PL group and 18.05 hrs in NS group, $P=0.5080$), but pH rise was more significant in PL group at 4 - 6 hrs and 6 - 12 hrs, and higher chloride levels and lower anionic gaps in NS group at 6 - 12 hrs. |
| 11 | DePiero et al.(39) | 1258 cases retrospective cohort study | Relationship between rehydration timing and complications | Children | - | Of the 1258 children with DKA, 27.8% developed hypertension, including 12.2% at the time of consultation and 15.6% during treatment. It is recommended that in the treatment of children with DKA, the decision to rehydrate should not be based on blood pressure alone, but should also focus on heart rate, peripheral perfusion, and other indicators. |
| 12 | Carrillo et al.(40) | 326 cases retrospective cohort study | The choice of rehydration type | Adults | LR vs. NS | The incidence of medical hyperchloremia was significantly lower in the LR group than in the NS group (64.2% vs 74.4%, $P = 0.05$), and the 48-hour serum creatinine decreased more significantly (-0.15 mg/dL vs -0.04 mg/dL, $P = 0.002$), which was more conducive to the recovery of renal function. |
| 13 | Poon et al.(41) | 556 cases retrospective cohort study | The choice of rehydration type | Children | - | Children in the DKA group were younger, had shorter duration of symptoms and some had infections. In terms of treatment, there were inconsistencies with international guidelines, such as the fact that only 58.9 per cent of children received saline shock rehydration in the emergency department and 12.4 per cent received insulin shocks. |
| 14 | Attokaran et al.(42) | 84 cases nested cohort study | The choice of rehydration type | Adults | PL vs. 0.9% NS | No significant advantage of PL in reducing ICU admission rates and length of stay was found. |
| 15 | Trainor et al.(43) | 753 cases retrospective cohort study | Relationship between rehydration timing and complications | Children | - | Accurate assessment of the degree of dehydration in children with diabetic ketoacidosis (DKA) is critical for rehydration therapy but is controversial and challenging. |
| 16 | Brown et al.(44) | 1277 cases retrospective cohort study | Determination of the amount and speed of fluid replacement | Children | - | Obese children and adolescents are more likely to receive rehydration below the protocol-specified rate during DKA therapy, yet higher rehydration rates based on body weight did not increase their risk of altered mental status or brain injury, and low rehydration rates increased the risk of hypophosphatemia. |
| 17 | Hay et al.(45) | 145 cases retrospective cohort study | Relationship between rehydration timing and complications | Children | - | 19% of children met the diagnostic criteria for AKI on admission and only 2.4% remained AKI on discharge. dehydration and high serum urea levels on admission were associated with high serum creatinine levels, fluid resuscitation and hyperchloremia were associated with delayed renal recovery, and the severity of AKI was associated with cerebral edema. The study suggests that the fluid regimen for DKA needs to be revisited to improve the prognosis of the child. |
| 18 | Johnson et al.(46) | 246 cases retrospective cohort study | The choice of rehydration type | Adults | NS vs. LR | The time to DKA remission (17.1 hours) and insulin drip duration (16.0 hours) were significantly shorter in the LR group than in the NS group (20.6 and 21.4 hours), and the LR group had lower serum chloride at 48 hours and a shorter ICU stay. |
| 19 | Jamison et al.(47) | 771 cases retrospective cohort study | The choice of rehydration type | Adults | LR vs. NS | The time to remission of high anion gap metabolic acidosis (HAGMA) was faster in the LR group (adjusted $HR=1.325$, 95% CI 1.121 - 1.566, $p<0.001$), and there were no differences between the two groups in terms of complication rates, length of insulin infusion, or length of hospitalisation. |
| 20 | Carter et al.(48) | 110 cases retrospective cohort study | The choice of rehydration type | Adults | BF vs. NS | Time to DKA remission was significantly shorter in the BF group (13 [10 - 19] hours) than in the NS group (17 [11 - 25] hours, $p = 0.02$). |

PL: plasma-lyte 148; NS: normal saline; LR: lactate Ringer's fluid; BF: equilibrium fluid; NA: not applicable.

### 4.3 Limitations

The limitations identified in this clinical trial include clinicians’ fluid resuscitation practices and the compliance of patients with diabetic ketoacidosis.

Some clinicians believe that larger volumes of fluid replacement yield better outcomes during the correction of acidosis. This concept permeates not only routine fluid therapy but also patient education. Consequently, some patients may perceive that reducing fluid volumes diminishes the effectiveness of rehydration, thereby hindering clinical trials.

Patients with DKA undergoing USCOM-guided fluid resuscitation face certain complexities compared to empirical fluid administration. We refer to USCOM-guided fluid resuscitation as titrated fluid therapy. During titrated fluid therapy, 250 ml of fluid is administered at each session, followed by combined USCOM and PLR measurements to determine whether fluid administration should continue. If fluid administration continues, another 250 ml is administered; otherwise, fluid administration is halted within 6 hours. This frequent assessment process may trigger negative emotions in patients, potentially compromising their compliance.

To mitigate these limitations, we conducted training for physicians administering emergency fluid therapy during the clinical trial to ensure that clinicians strictly adhered to the study protocol. Additionally, we patiently explained the complex procedures involved in the trial when participants signed their informed consent forms. Participants could withdraw from the clinical trial at any time if they expressed dissatisfaction or refusal to accept the titration-based fluid therapy protocol after enrollment. These two measures maximized the mitigation of the limitations.

### 4.4 Generalisability

This study demonstrates for the first time that USCOM has significant advantages in guiding fluid resuscitation in DKA patients, which can effectively reduce fluid overload and shorten the length of hospital stay without compromising metabolic recovery and glycaemic control.

We recommend that clinicians in other countries and regions adopt our principles and methodology for titrated fluid resuscitation, then implement them in ways that align with local policies and healthcare environments. For instance, patients receiving fluid therapy alone without management of complications can still benefit from USCOM-guided resuscitation. This approach allows the dissemination of new research findings globally while mitigating regional and population-specific variations.

### 4.5 Fluid management implications

Achieving rapid, voluminous, yet safe fluid resuscitation remains a fundamental challenge in DKA management. This pilot trial introduces a novel solution: dynamic fluid responsiveness assessment via USCOM integrated with PLR. Before each fluid bolus, patients underwent USCOM-PLR evaluation; only fluid responders (ΔSV ≥10%) received additional crystalloid. This protocol objectively identifies responders, overcoming the limitations of empirical formulas and significantly mitigates fluid overload risk while ensuring adequate resuscitation.

We elaborate on the clinical translation implications of our findings from two perspectives: patient benefits and insights into clinicians’ thinking.

Patients with diabetic ketoacidosis (DKA) face two key treatment challenges during fluid therapy: correcting acidosis and managing complications. In the United States, the average hospital stay for DKA fluid therapy is 3.5 days. This duration solely accounts for the time required to correct acidosis. This may be linked to the country’s high healthcare costs, which can make it financially prohibitive for patients to cover the expenses associated with DKA complications. In other words, DKA patients must rely on home care and self-management to address complications. In China, the cost of correcting acidosis is approximately 500 RMB, while managing complications typically ranges from 5,000 to 15,000 RMB depending on complexity (data from the Emergency Department of Yangzhou University Affiliated Hospital). According to the 2025 Yangzhou Municipal Government report, the minimum monthly wage for Yangzhou residents is 2,490 RMB. This indicates that while correcting acidosis is affordable for the average resident, managing complications imposes at least a tenfold increase in medical burden. Our findings show that patients using USCOM-guided fluid replacement experience an average increase of 952.56 ml in urine output during the first 24-hour infusion period. High urine output stimulates fluid-sensitive loading in DKA patients, directly alleviating cardiac volume overload and reducing heart failure risk. This protective mechanism prevents fluid-related complications. For patients in China, this mechanism shortens the average hospital stay by 1.92 days after additional complication management. Thus, USCOM-guided fluid therapy enables patients to endure lower medical costs during complication management while benefiting from shorter hospital stays.

Additionally, in the context of training young clinicians in fluid resuscitation skills, using USCOM guidance provides objective evidence to help them master these skills compared to empirical fluid administration. Once the patient has the USCOM in place, fluid therapy can be initiated. Each infusion delivers 250ml. Subsequently, read the USCOM parameters and use the PLR to determine whether to continue fluid administration. This strategy is termed titrated fluid therapy. If ΔSV < 10, it indicates fluid administration cannot continue, as the patient has reached fluid overload. Repeat the above steps every 6 hours thereafter. The USCOM-guided fluid administration process not only helps young clinicians master fluid management skills but also objectively controls fluid volume.

During the titrated fluid replacement process, we observed an intriguing phenomenon. Although patients received a greater volume of fluids, the final balanced fluid volume was relatively reduced. We attempted to explain this phenomenon. The fundamental reason DKA patients require massive fluid replacement lies in severe fluid loss causing systemic pathophysiological changes; fluid replacement is the first and most critical step to reverse these changes. The core mechanism involves hyperglycemia inducing osmotic diuresis, which exacerbates dehydration and electrolyte depletion, perpetuating a vicious cycle of worsening acidosis. Regardless of the underlying cause, sharply elevated blood glucose levels exceed the kidneys’ reabsorption capacity, resulting in massive glucose excretion in urine. As an osmotically active substance, glucose “grabs” water for excretion, significantly increasing urine output. This osmotic diuresis causes substantial fluid loss. Reduced circulating blood volume leads to decreased blood pressure, tachycardia, inadequate tissue perfusion, and even shock. Concurrent with massive water loss, electrolytes (particularly sodium, potassium, magnesium, and phosphate) are also lost in large quantities. Severe dehydration and hypovolemia cause insufficient tissue perfusion, triggering lactic acid accumulation and worsening metabolic acidosis. Circulatory failure also diminishes the kidneys’ capacity to excrete acids. Thus, the primary physiological goals of aggressive fluid resuscitation are: Restore effective circulating volume to improve tissue perfusion and correct shock. Dilute blood glucose and plasma osmolality, creating an environment conducive to insulin action. Enhance renal perfusion to facilitate clearance of excess glucose, ketones, and acidic substances. In essence, fluid resuscitation “refills” the vascular and interstitial spaces depleted by hyperglycemia, serving as the cornerstone for breaking the vicious cycle of DKA. Patients receiving USCOM-guided care initially received higher fluid volumes, likely due to assessments of volume responsiveness. Physicians deemed titrated therapy to be more evidence-based, enabling more aggressive and decisive early fluid administration. This early aggressive fluid administration rapidly corrects the vicious cycle of fluid loss, improving renal perfusion, increasing urine output, and reducing net fluid intake. The decrease in net fluid intake may also stem from titrated therapy, which minimizes the risk of overhydration associated with empirical fluid administration.

## 5 Conclusion

Uscom-guided fluid management is a clinically translatable strategy for balancing the efficacy and safety of DKA resuscitation, especially in resource-limited settings where advanced monitoring is lacking.

## Data Availability

All data produced in the present study are available upon reasonable request to the authors.

## Acknowledgments

The authors thank Yangzhou University Affiliated Hospital for technical assistance. The study complied with the Declaration of Helsinki and received approval from Yangzhou University Affiliated Hospital Review Board (IRB-06-004). All participants provided written informed consent prior to enrollment. The protocol was registered with the Chinese Clinical Trial Registry (ChiCTR2500103388) and conducted in accordance with CONSORT guidelines. The use, analysis and dissemination of this clinical research data were permitted by Prof. Dr. Yifei Chen of the Affiliated Hospital of Yangzhou University.

## Generative AI statement

The authors declare that no Gen AI was used in the creation of this manuscript.

