## Supplementary document for "Ultrasonic Cardiac Output Monitor-Guided Fluid Resuscitation in Adult Diabetic Ketoacidosis: A Randomised Controlled Trial"

**Running Title: A Randomised Controlled Trial**

**Yan Bo**[1*]

[1]The Department of Medicine, Northwest Minzu University, Lanzhou 730000, Gansu Province, China

***Correspondence:**

Yan Bo

### Supplementary document

#### Supplementary document Table 1. Outcome

| Outcome | Admission | First fluid resuscitation | Day 1 | Day 2 | Day 3 | Last fluid resuscitation | Day 4 | … | Day 13 | Day 14 | Discharge |
|---|---|---|---|---|---|---|---|---|---|---|---|
| Fluid replacement | | | ✓# | ✓# | ✓# | | ✓ | ✓ | ✓ | ✓ | |
| Days of hospitalisation | | | | | | | | | | | ✓# |
| Complications | | ✓ | ✓ | ✓ | ✓ | ✓ | ✓ | ✓ | ✓ | ✓ | ✓# |
| Time to urine ketone conversion | | | ✓ | ✓ | ✓ | | ✓ | ✓ | ✓ | ✓ | ✓# |
| Blood glucose | ✓# | | ✓# | ✓ | ✓# | | ✓ | ✓ | ✓ | ✓ | ✓ |
| USCOM technical parameters | | ✓# | ✓ | ✓ | ✓ | ✓# | | | | | |

✓ indicates the time point of the measurement in the study. # indicates that the data measured at that time point are reported in the results.

#### Supplementary document Table 2. Baseline Demographic and Clinical Characteristics

| Parameter | USCOM Group (n=25) | Control Group (n=25) | Statistical effect size | P-value |
|---|---|---|---|---|
| Age (years) [a] | 51.8 ± 13.5 | 44.1 ± 18.1 | 1.70 | 0.093 |
| BMI (kg/m²) [a] | 24.1 ± 2.5 | 24.8 ± 4.3 | -0.74 | 0.464 |
| SBP (mmHg) [a] | 127.1 ± 16.1 | 132.5 ± 23.1 | -0.95 | 0.346 |
| DBP (mmHg) [a] | 77.8 ± 10.6 | 76.0 ± 15.5 | 0.47 | 0.641 |
| MAP (mmHg) [a] | 94.2 ± 11.5 | 94.8 ± 16.8 | -0.15 | 0.881 |
| Heart Rate (bpm) [a] | 102.2 ± 19.4 | 104.1 ± 18.1 | -0.35 | 0.731 |
| Respiratory Rate (breaths/min) [a] | 24.7 ± 6.1 | 22.5 ± 5.1 | 1.35 | 0.180 |
| $SpO_2$ (%) [a] | 96.6 ± 2.1 | 96.2 ± 1.4 | 0.80 | 0.431 |
| Sex (Male/Female) [b] | 11/14 | 10/15 | 0.04 | 0.396 |

a: t-test; b: Pearson's $\chi^2$ test. P-value is the result of two-sided test, and $p<0.05$ indicates that the difference is statistically significant. BMI: body mass index; SBP: systolic blood pressure; DBP: diastolic blood pressure; MAP: mean arterial pressure; $SpO_2$: peripheral capillary oxygen saturation.


This is a provisional file, not the final typeset article

**Supplementary document Table 3. Incidence of complications in USCOM-Guided vs. Control Groups**

| Complication | USCOM Group (n=25) | Control Group (n=25) | $\chi^2$ | Cohen's d | Absolute Risk Difference | 95%CI | P-value |
|---|---|---|---|---|---|---|---|
| Hypoglycemia [a] | 6 (24%) | 5 (20%) | 0.12 | 0.086 | +0.04 | (−0.19, 0.27) | 0.733 |
| Hypokalemia [a] | 6 (24%) | 5 (20%) | 0.12 | 0.086 | +0.04 | (−0.19, 0.27) | 0.733 |
| Pulmonary Edema [b] | 0 (0%) | 1 (4%) | – | 0.252 | -0.04 | (−0.12, 0.04) | 0.313 |
| Acute Kidney Injury [b] | 2 (8%) | 3 (12%) | – | 0.134 | -0.04 | (-0.21, 0.09) | 0.640 |
| Arrhythmia [b] | 5 (20%) | 3 (12%) | – | 0.274 | +0.08 | (−0.12, 0.28) | 0.440 |

a: Pearson's $\chi^2$ test; b: Fisher's exact test. 95% CI: 95% confidence intervals.

**Supplementary document Figure 1. Flow diagram.** Flow diagram of the progress through the phases of a randomised trial of two groups drawn by Figdraw.

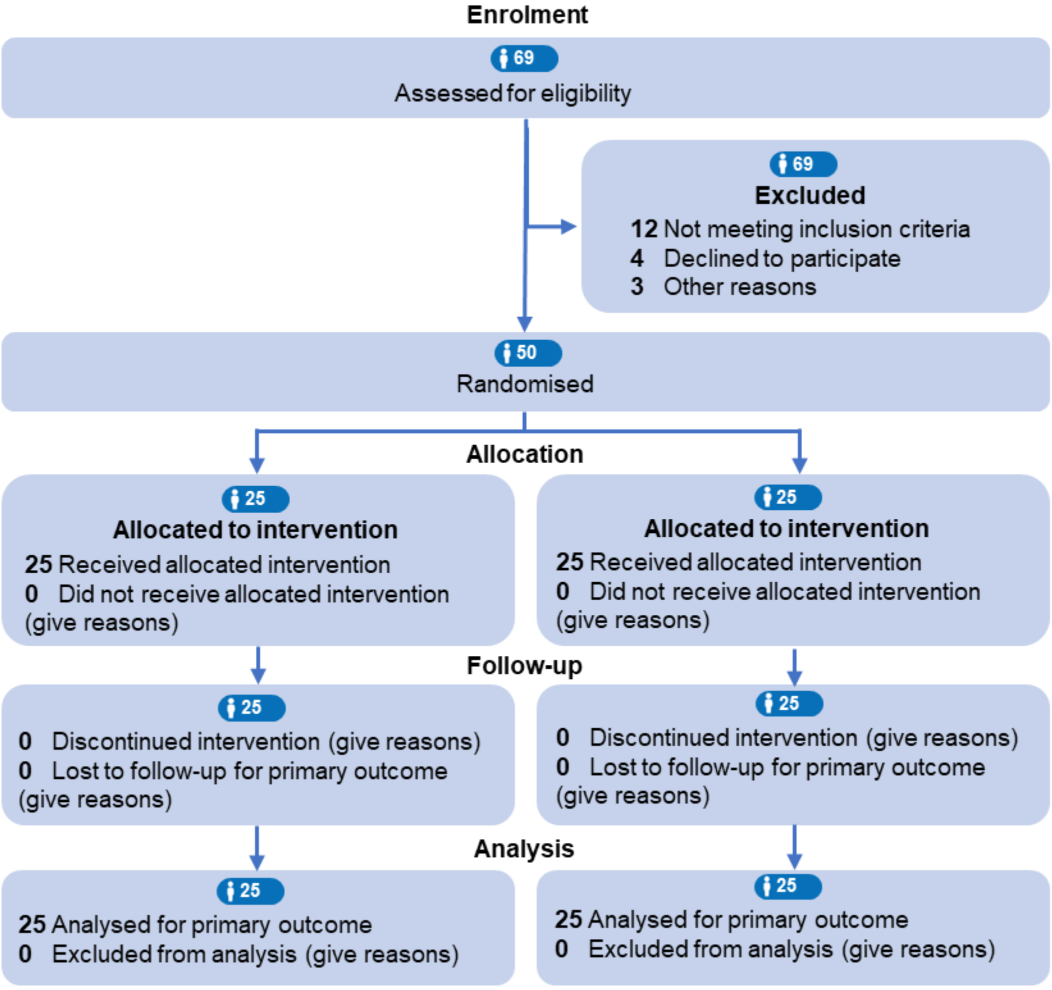
